# Bagged Fuzzy-Rough Nearest Neighbors (BFRNNs): A Novel Ensemble Learning Algorithm for Disease Diagnosis and Prognosis Prediction

**DOI:** 10.1101/2023.10.21.23297353

**Authors:** Aashish Cheruvu, Lockheed Martin

## Abstract

Purpose of the study is to develop a novel machine learning (ML) algorithm that can accurately predict malignant versus benign tumors. A novel ML hybrid ensemble model called “Bagged Fuzzy-Rough k-Nearest Neighbors” (BFRNN) was developed. BFRNN is an improvement over the widely used k-Nearest Neighbors algorithm due to its use of fuzzy-rough logic and an unique ensemble voting algorithm. Initially, graphical libraries were used to visualize the Wisconsin Breast Cancer biomarker dataset (WBCBD) to capture useful insights about the data. Following preprocessing of the data (e.g. encoding categorical data snd removing outliers), a small subset of the most important breast cancer biomarkers were chosen based on feature selection technique and applying breast cancer domain knowledge. The performance of BFRNN was compared with a sample of five commonly used ML classification algorithms. The criteria for the evaluation the performance of ML was based on accuracy, area under the Receiver Operating Characteristic curve, and the ability to overcome overfitting. Discussion: Among the algorithms evaluated, BFRNN was the best classifier of WBCBD achieving an average training score of 98.47% and an average testing score of 99.09%. Among the other common ML algorithms evaluated, the highest test accuracy observed was 95.1% for Random Forest, with significant overfitting. In addition, outlier removal from the dataset and Pearson’s Correlation evaluation steps can be avoided for the implementation of the BFRNN algorithm. BFRNN has shown high accuracy in classifying the malignant versus benign characteristics and this algorithm could be a useful tool in disease diagnosis.

## I. Introduction

With the ever-growing list of novel diagnostic tools in the fight against cancer, there is an exponential growth in the number of biomarkers (e.g laboratory, imaging markers and tumor molecular profiling) that are used in the diagnosis. With the availability of these vast amounts of data, cancer diagnosis based on the knowledge acquired through experience and self-learning is challenging. To this end Machine Learning (ML) methods that can process huge amounts of data in multiple dimensions are the most promising tool to assisting physicians. The algorithms used in ML have the ability to discover, classify, and identify patterns and relationships between various disease characteristics and effectively predict future outcomes of disease. Breast cancer is the second most common cancer among women in the United States [1]. In the diagnosis of the breast cancer, physicians use various biomarkers like Estrogen Receptor (ER), Progesterone Receptor (PR), Human epidermal growth factor receptor 2 (HER2) etc. Among the various breast cancer biomarkers, Wisconsin Breast Cancer biomarker dataset (WBCBD) has been used extensively in the literature [2]. Some of the most common algorithms studied were K-nearest neighbor, logistic regression, Decision trees, Neural Networks, Naïve Bayes classifier, Support vector machine, and Deep Learning model etc. Each algorithm is used to construct a classifier model using training dataset [3]. Based on testing and performance measure the classifier model will be used for classifying unknown instances. The current study evaluated a novel model using k nearest neighbors (kNN) and Fuzzy-Rough set theory. The method was ensembled with bootstrap aggregating (also known as bagging). This work differs from other publication regarding the application of fuzzy-rough theory as a classifier of medical data. Firstly, the BFRNN model utilizes ensembling methods to reduce variability during model evaluation. Secondly, the model makes use of unique voting methods in order to improve the evaluation metrics. In the present study, the baseline predictive algorithms like kNN, logistic regression, random forest, linear support vector classifier, deep learning algorithms were also evaluated for comparative evaluation. The objective of this work is to develop a novel machine learning algorithm that can applied in healthcare, accurately diagnosing the presence of the breast cancer by utilizing structured biomarker data.

## II. Materials

In this paper we have used Wisconsin Breast Cancer Dataset that are publicly available for researcher this database is generated from biopsy images that having 569 rows and 30 columns of dataset [4]. The programming platform used was Colaboratory, a cloud Service made by Google that allows Python to run the code in a notebook format and access GPUs and TPUs to speed up the runtime of programs and allows for easy sharing of code. The programming language used was Python. For data visualization matplotlib and Seaborn were used. Machine learning libraries used were NumPy, pandas, sci-kit learn, Tensorflow, Keras.

## III. Methodology

The machine learning workflow is illustrated in Figure 1.

**Fig. 1:**
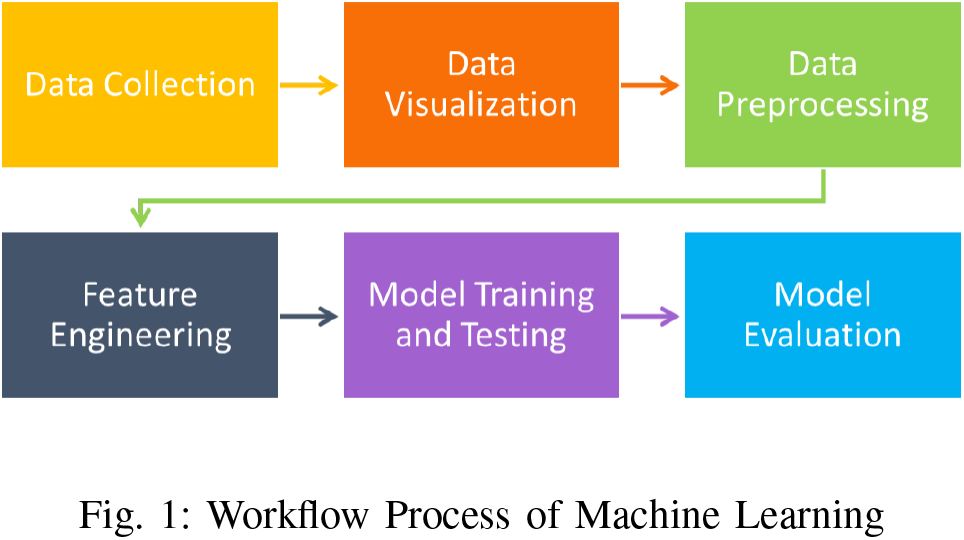
Workflow Process of Machine Learning

### A. Gathering Data

As shown in Figure 1, the first step involved in the machine learning workflow was gathering data. In the current analysis, Wisconsin Breast Cancer biomarker dataset (WBCBD) created by Wolberg and contributed by other researchers of the University of Wisconsin was used. This dataset was constructed from various imaging biomarkers calculated from a fine needle aspirate taken from subjects with and without breast cancer. The data set consists of data from 569 subjects of which 357 subjects have benign and 212 are malignant tumors with 30 biomarker features.

### B. Data Visualization

The second step in the machine learning workflow was data visualization. Data visualization helps translating data into a visual context, such as a map or graph. The main goal of data visualization is to make it easier to identify patterns, trends and outliers in large data sets. The data visualization in the current analysis was performed using matplotlib and seaborn. The forms of data visualization created are i) distribution plots, ii) heatmaps, ii) pair plots, iii) kernel density estimate plots and iv) boxplots. The data visualization along with the data preprocessing helped in identifying and shortlisting the biomarkers that are informative about the cancer status.

### C. Data Preprocessing

The third step of the machine learning workflow was the preprocessing of data. WBCBD is a well cleaned dataset with no missing values. For the data preprocessing, the data required one-hot encoding and outlier removal.

#### 1) One-hot encoding

One-hot encoding forcing a categorical variable to be a numerical (e.g. benign vs malignant), was assigned on the target variable (whether the subject was reported as having or not having cancer).

#### 2) Outlier Identification

An outlier is an observation that lies outside the overall pattern of a distribution [5]. Outliers can adversely affect the training process of a machine learning algorithm, resulting in a loss of accuracy. The outliers in the WBCBD were identified using Inter Quartile Range method and Density-based spatial clustering of applications with noise (DBSCAN) method. DBSCAN is an unsupervised density-based clustering method which is used for separating clusters of high density from clusters of low density. DBSCAN divides the dataset into multiple dimensions. Then, it picks a random point and calculates the number of nearby points. It continues the process until no other data points are nearby, and then it will look to form the second cluster. Two points are said to be in the same cluster if one-point falls within the radius (epsilon) distance of another. Furthermore, the minimum number of points needed is another parameter which determines the clusters. While going through each data point, a cluster is formed when the DBSCAN finds the minimum number of points needed within epsilon distance of each other. The point which is having the minimum number of points needed within epsilon distance is called as ‘core point’. hence, the points which do not fall under any cluster are considered to be outliers. The data preprocessing, outlier identification and machine learning algorithm methods used in the current analysis is outlined in Figure 2.

**Fig. 2:**
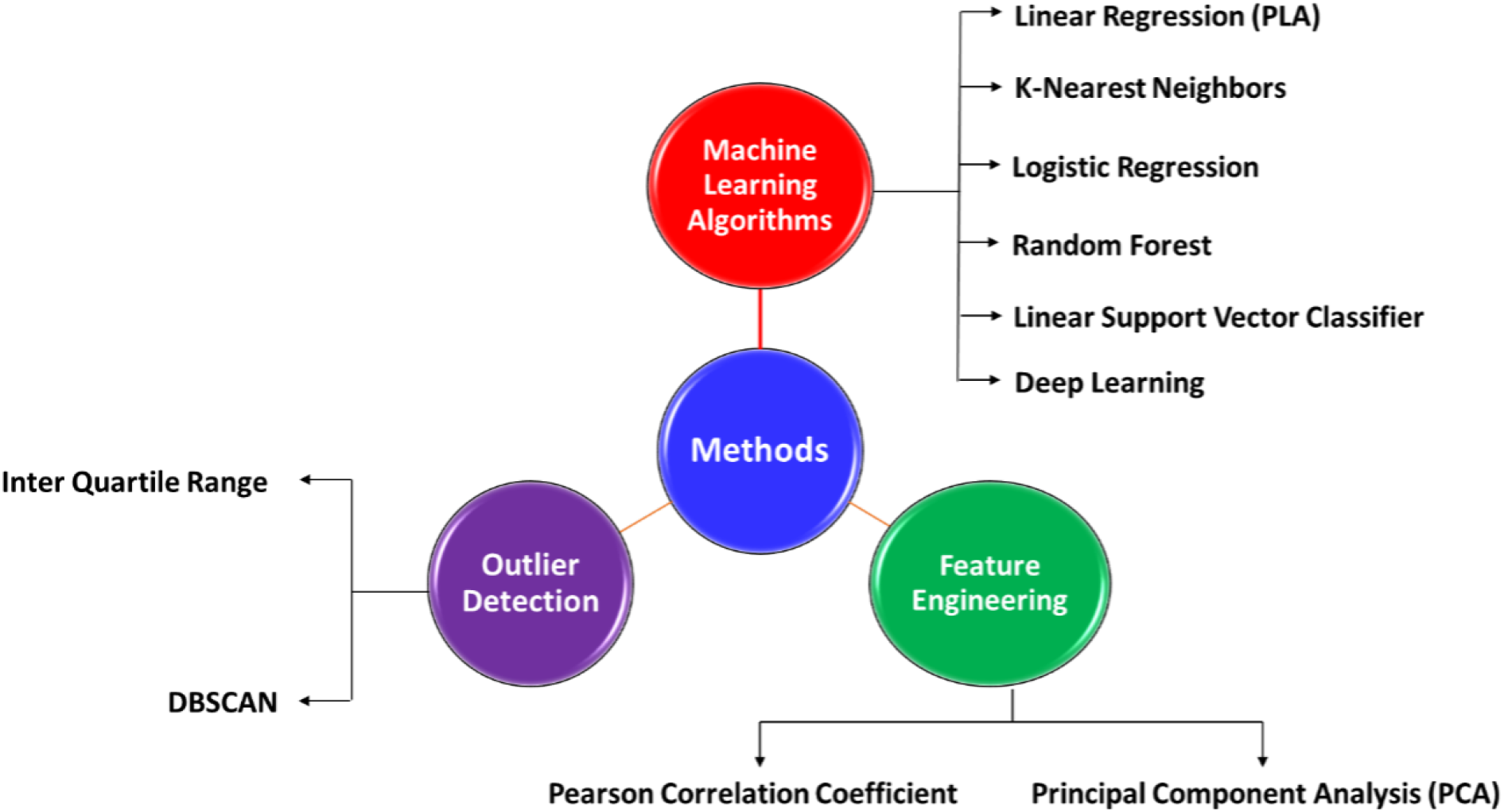
Summary of Methods Used in Developing ML algorithms^1^

### D. Feature Engineering

Feature engineering is the process of transforming raw data into features that better represent the underlying problem to the predictive models, resulting in improved model accuracy on unseen data. Feature selection and feature extraction methods were performed on the data to reduce the dimension of features, thereby producing reduced versions of the original dataset. With the current data set two feature engineering techniques were applied: Pearson’s correlation coefficient and Linear Discriminant Analysis.

#### 1) Pearson’s correlation Coefficient

A Pearson correlation is a number between -1 and 1 that indicates the extent to which two variables are linearly related. The correlation coefficient has values between -1 to 1. A value closer to 0 implies weaker correlation (exact 0 implying no correlation). A value closer to 1 implies stronger positive correlation. A value closer to -1 implies stronger negative correlation.

#### 2) Linear Discriminant Analysis

Linear discriminant analysis (LDA) is one of the statistical feature extraction methods used to reduce high dimensions of data. It is mainly used for supervised dimension reduction. This means that it takes into consideration the different class labels. LDA is a feature extraction technique that computes transformation by maximizing the between class scatter and minimizing the within class scatter. This is done simultaneously, and the highest-class discrimination is achieved. To compute the optimal transformation in LDA, Eigen decomposition is used on the co-variance matrices.

### E. Machine Learning Algorithms

The ML algorithms used in the classification of benign and malignant were Linear Regression (PLA), K-Nearest Neighbors (KNN), logistic regression, random forest, linear support vector classifier and deep learning techniques.

#### 1) KNN

The term KNN means K nearest neighbor, a popularly used simple classification machine learning algorithm. It uses information about a tuple’s K nearest neighbors to classify unlabeled tuples. The value of K can be any number. After deciding K, the algorithm requires a training dataset to be classified into classes as labelled by a target class variable. Then, for each unlabeled tuple in the test dataset, KNN identifies the “K” nearest neighbors and assigns a new observed data point to the class most present among the “neighbors”.

#### 2) Logistic Regression

Logistic regression seeks to provide a nonlinear decision boundary in the form of a sigmoid curve (also known as a logistic curve, hence the name) defined by the equation 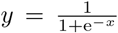. Logistic regression takes data points and calculates the sigmoid value. The discrimination of classes is performed by comparing the sigmoid value to a set threshold (usually 0.5) above and below which the data points are assigned to different classes.

#### 3) Random Forest

Random Forest classifier is an ensemble learning model (multiple classifiers are combined together to form predictions) created by combining many decision trees (yes-no decisions are made at different branches, and the final prediction of the model based on the yes-no decisions are called leaves) by the use of a technique called bagging, or bootstrap aggregating. Bootstrap aggregating is a process in which different models (in this case, decision trees) are trained via bootstrapped data sets. Each model makes a vote for what class the data point should be classified as, and the data point will be classified based on the majority decision.

#### 4) Linear Support Vector Classifier

A Support Vector Machine (SVM) is a hyper plane that creates boundaries between points of data plotted which represent tuples and their feature values. The SVM learner combines the aspects of both the KNN model of learning and the linear regression model. The combination is highly robust, making SVM able to model highly complex problems.

#### 5) Multilayer Perceptron

Neural networks consists of layers of neurons subclassified into three categories: input layer neurons, hidden layer neurons, and output layer neurons. Input layer neurons take in the data and pass it to the hidden layers, which calculate a weighted average calculation of the different features passed into the input neurons. The weights and biases are updated again and again through a process called backpropagation until the model converges, finding the optimal weights and biases. The output neuron layer then calculates and classifies each data point.

#### 6) Bagged Fuzzy-Rough Nearest Neighbors

Bagged Fuzzy-Rough Nearest Neighbors (BFRNN) is an novel ensemble model that was developed based on the fuzzy-rough set. The fuzzy set [6] is a set where all data points are considered members of a certain class to an extent – each data point is not discretely classified as benign or malignant, but rather classified on a continuous scale where points are considered 20 or 30% malignant. These fuzzy sets are then taken as parameters for the rough set (a set which takes upper and lower approximations of other sets to make a decision boundary that is not definite but rather a range) in order to make the decision boundaries of ML algorithms more “fuzzier” and much less defined. This fuzzy-rough set approach was then applied to the k-Nearest Neighbors classifier. The result combination – fuzzy-rough nearest neighbors’ classifier, or FRNN, has been shown to work well in medical data classification. However, since fuzzy logic classifies points on a spectrum, the FRNN algorithm tends to be variable in its accuracy at classifying data points (because the classification of points can change constantly. e.g. a point’s classification changes from 0.64 to 0.67). To make the classification of data points less variable, a bagging algorithm [7] was applied (multiple FRNN classifiers are set up and vote on the final class of the algorithm, making each individual FRNN’s variable predictions less contributive towards the final classification). However, literature has shown that standard bagging on a k-Nearest Neighbors based classifier is at best no help improving accuracy, and at worst, can decrease the classification accuracy. However, by implementing a unique voting technique, rather than simple majority voting, bagging can improve model accuracy. Therefore, a unique voting method that uses Borda Count (a weighted voting method, in which certain votes are given more importance than others) [8] was applied to the bagging algorithm. Each FRNN uses Borda Count to classify data points, where neighbors of data points that are closer to the data point to be classified are given more weight in the final decision than farther neighbors. Then, after each algorithm had come with a new prediction, the class with the greatest number of predictions among all of the classifiers (simple majority vote) will be the overall BFRNN’s prediction of the class of the data point. This unique voting method is known as Majority Component Classifier Borda Count. Further, a specific type of feature selection, known as decision tree information gain was used as for feature selection, due to its good integration with bagging algorithms (Random Forest is a popularly used example of a model that uses Information Gain and bagging). The schematic used in the novel BFRNN classifier was shown below in Figure 3.

**Fig. 3:**
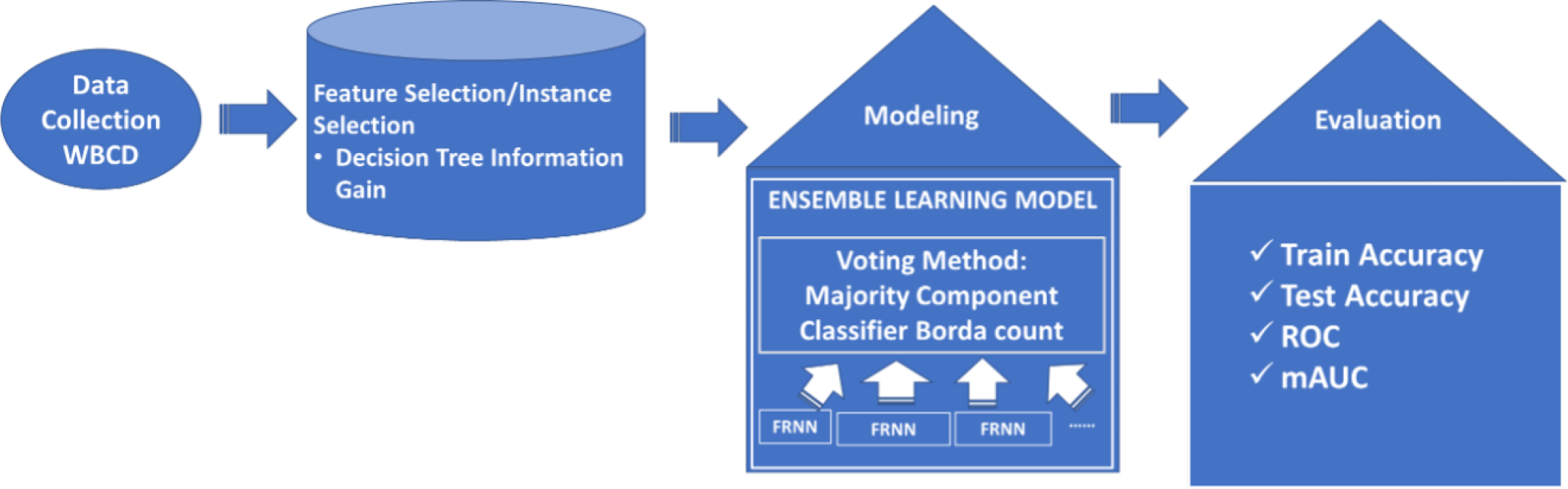
Schematic used with Bagged Fuzzy-Rough Nearest Neighbors (BFRNN)

## IV. Results and Discussion

### A. Data Visualization

#### 1) Distribution Plots

Figure 4 shows the distribution plots with histograms for each biomarker. As shown in the figure some biomarkers, such as symmetry error have high overlap between malignant and benign tumors, making it less informative for the algorithm to train on. However, with other biomarkers such as worst perimeter, are very reliable at classifying malignant vs. benign. It can also be observed that, generally, the spread of biomarker data of subjects with malignant tumor is much larger than the spread of benign biomarker data of subjects without cancer.

**Fig. 4:**
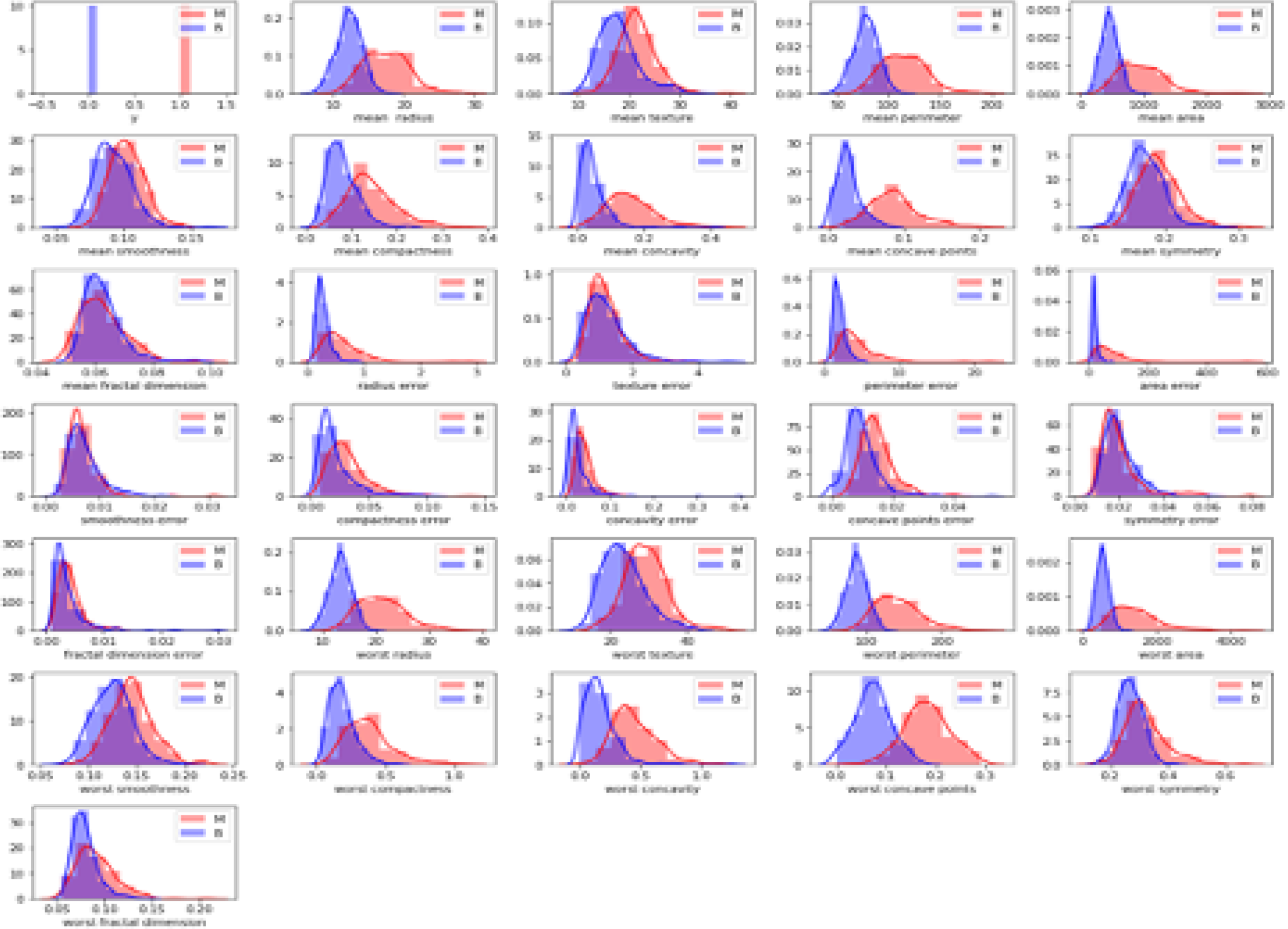
Distribution plots for all the biomarkers

### B. Data Preprocessing

#### 1) One-hot encoding

One-hot encoding was applied to the data set, by encoding the categorical variable of benign tumors to 0 and the malignant tumors to 1.

#### 2) Outlier Detection “Inter Quartile Range” Method

Data outliers were identified using the data points outside the 1.5 * IQR range. The box plot with the outliers for the shortlisted eight biomarkers are included below in Figure 5. The outliers identified using the IQR were only omitted from the baseline prediction algorithm evaluations.

**Fig. 5:**
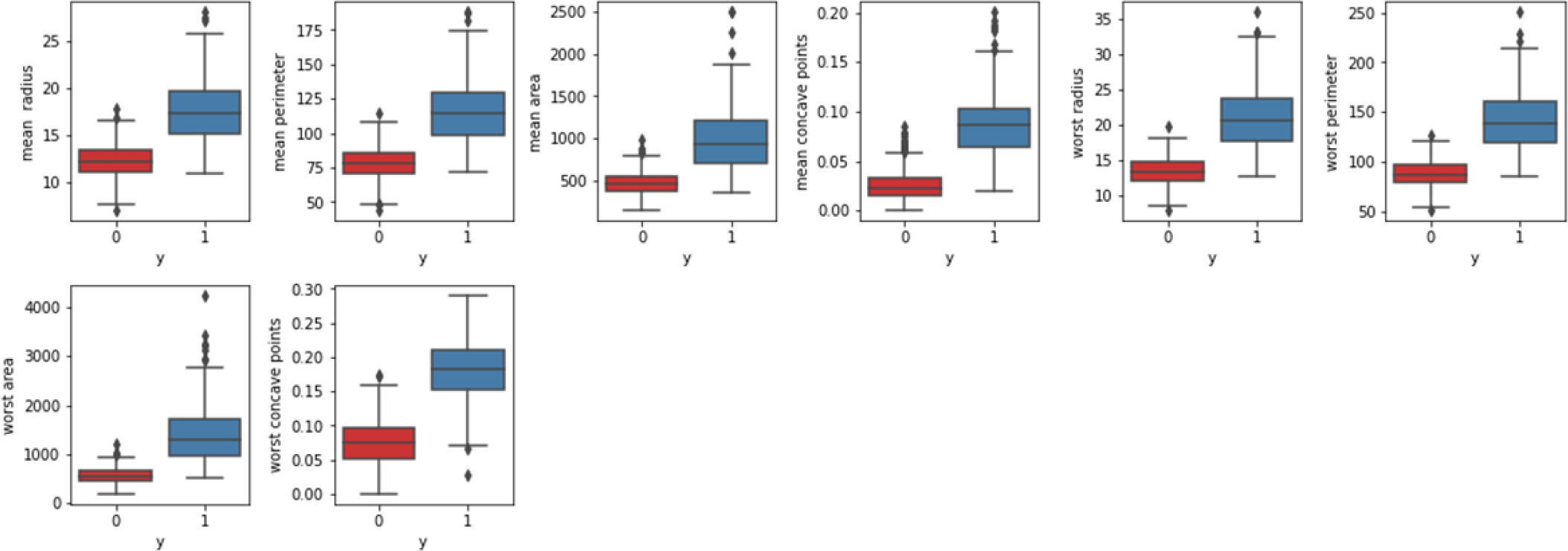
Box plots of the eight key biomarkers

### C. Feature Engineering

#### 1) Heatmaps

Figure 6 shows the heatmap of the Pearson’s Correlation coefficients (PCC) between all 30 biomarkers. In addition, the column labelled “y” shows the PCC for each of the biomarkers’ ability in predicting whether a tumor is benign or malignant. The color of the individual cells represents how strong the correlation is between the biomarkers. The warmer the color, the higher the correlation between the biomarkers – a blue cell represents a low correlation (below 0.5), and a red cell represents a high correlation (above 0.5). Based on the PCC scores, eight key features were found to have the correlation of coefficient of 0.7 or greater.

**Fig. 6:**
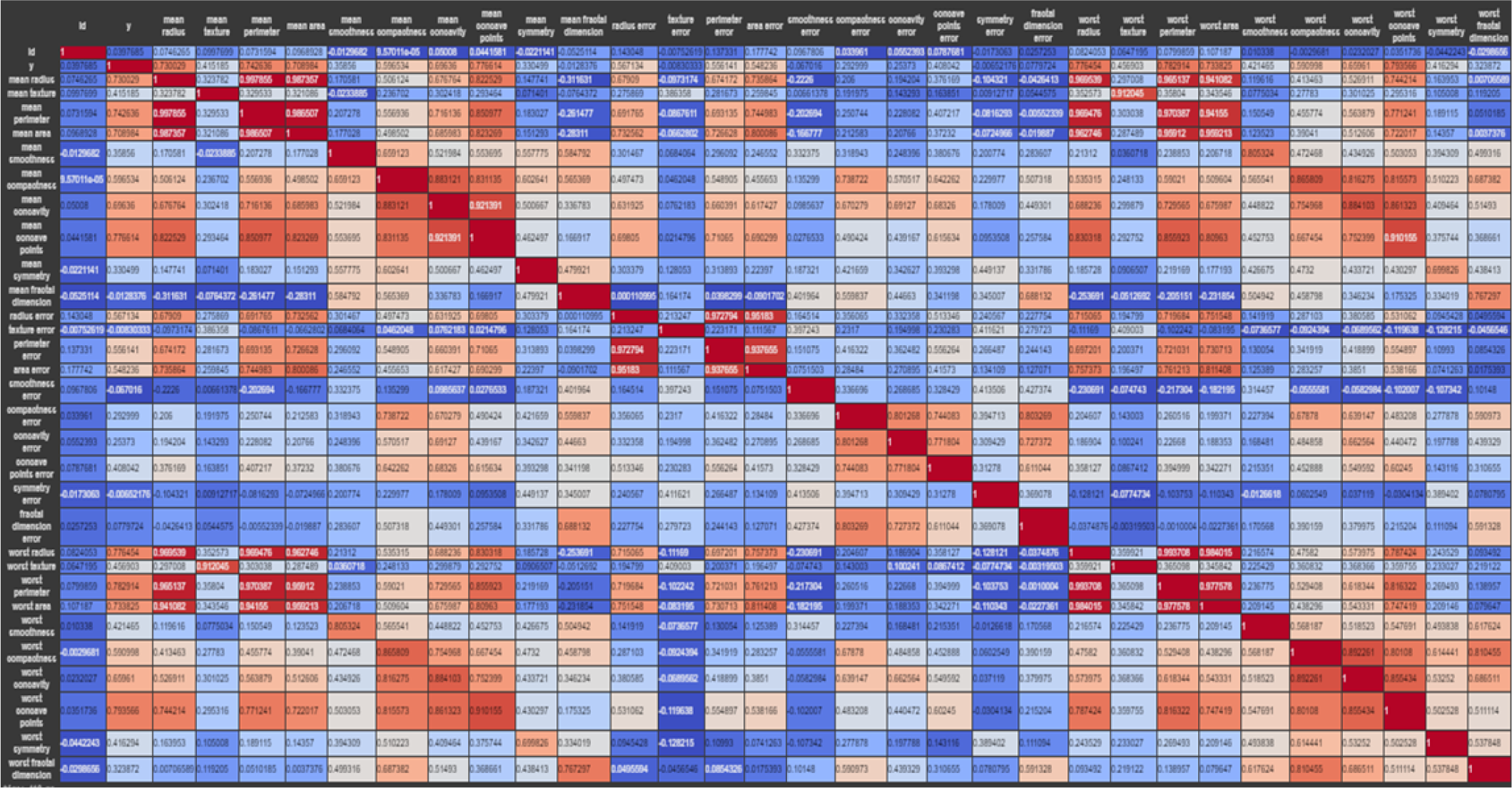
Heatmap of all biomarkers

Figure 7 shows a heatmap of the biomarkers that are best at classifying a tumor as benign or malignant. As shown in Figure 7 the level of correlation of each biomarker with the column labelled “y” is greater than 70% for all 8 biomarkers.

**Fig. 7:**
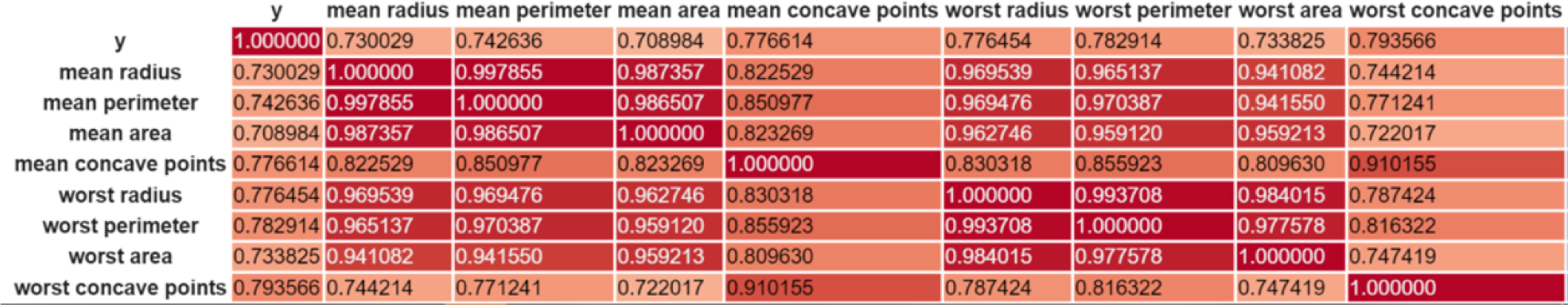
Heatmap of the key biomarkers

#### 2) Pair Plots

Using the eight selected biomarkers, based on PCC, Pair Plots for the 8 selected biomarker features were created. As shown in Figure 8, while the eight biomarker features seem mostly separable, there is a strong correlation observed between the biomarker features of “worst perimeter” and “worst radius” and “mean perimeter” and “mean radius”. When looking back at how the features were collected, it was determined that the radius features were derived from the perimeter features.

**Fig. 8:**
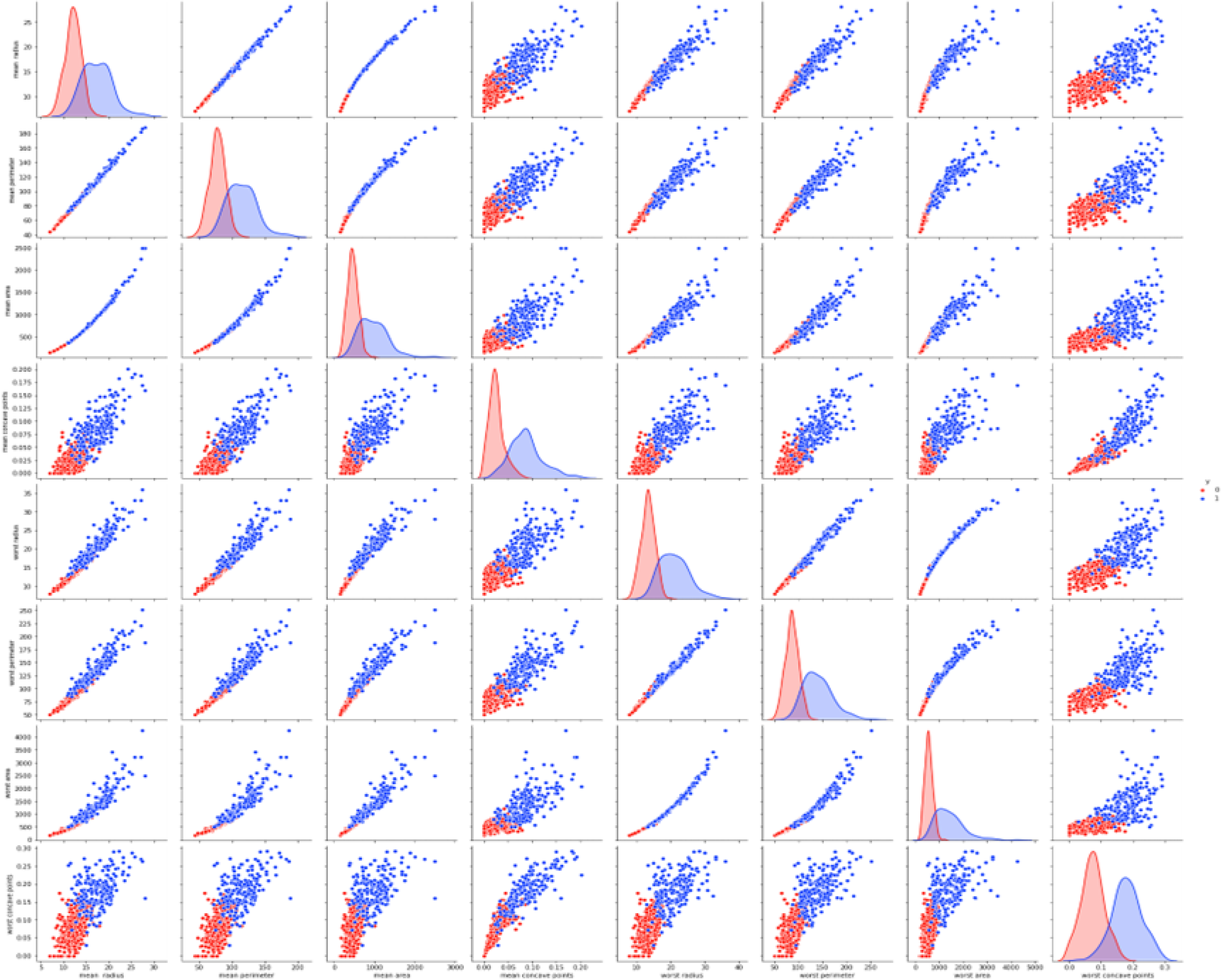
Pair plots of the eight key biomarkers

#### 3) Density-based spatial clustering of applications with noise (DBSCAN)

The DBSCAN plot for all the 30 biomarkers and the eight selected biomarkers was included below in Figure 9. The variables used to plot the DBSCAN graph were the first and second principal components. The DBSCAN output in Figure 9 shows how the data is more clustered and less dispersed following feature engineering.

**Fig. 9:**
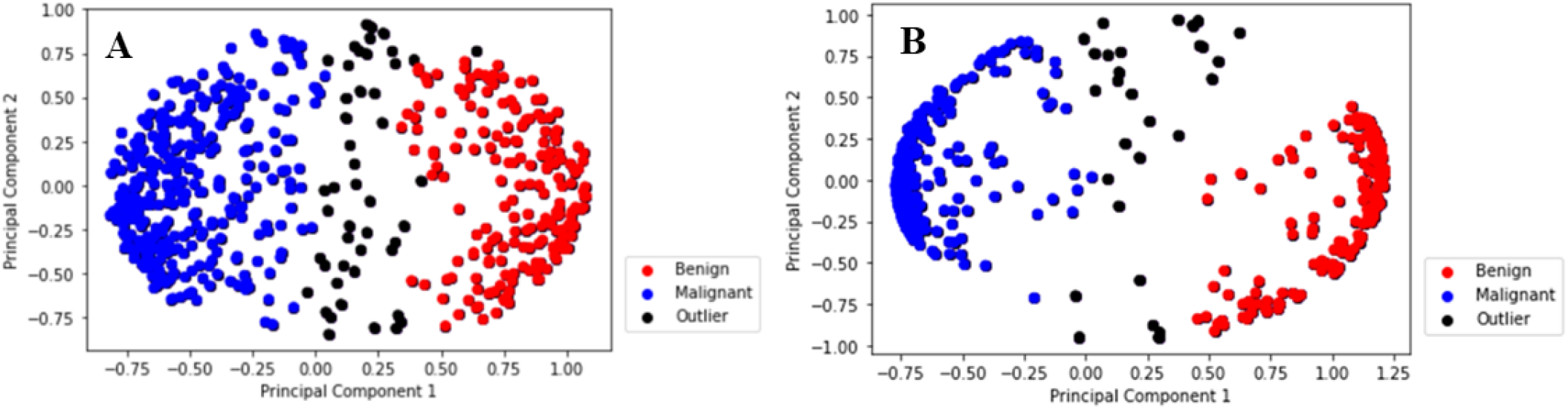
Comparative DBSCAN plots of the A) all biomarkers and B) the key biomarkers

### D. Machine Learning Algorithms

The summary of the machine learning algorithms performance is included in Table I. Each of the algorithms were run 10 times and the average training, testing and mAUC scores were reported.

**TABLE I:** Performance of Various Algorithms.

| Algorithm | Average Training Score (%) | Average Testing Score (%) | Measured Area Under the Curve (mAUC) (%) |
| --- | --- | --- | --- |
| K-Nearest Neighbors | 96.4 | 94.5 | 0.97 |
| Logistic Regression | 95.98 | 94.4 | 0.982 |
| Random Forest | 98.53 | 95.42 | 0.978 |
| Linear Support Vector Classifier | 95.7 | 95.8 | 0.987 |
| Deep Learning | 96.09 | 95.275 | 0.9858 |
| Bagged Fuzzy-Rough k-Nearest Neighbors Classifier | 98.475 | 99.091 | 1.0 |

#### 1) k-Nearest Neighbors Algorithm

k-Nearest Neighbors performed well, considering how it classifies points as malignant or benign, discussed in the Methodology section of this research paper. The k-NN algorithm performed the well with a training and testing accuracy of 96.4% and 94.5%, respectively. Due to how k-NN works, it can be noted that the WBCBD ‘s FNA biomarker values are strongly clustered around data points that are similarly classified. Due to the strength of k Nearest Neighbors at classifying malignancy, it was thought that combining this classifier with a modern technique called the fuzzy rough set and a bagging technique would improve the performance of the model at classifying breast cancer, and the results of BFRNN show this thought was successful. More generally, across all the baseline predictive models, the mAUC scores were very similar, leading us to believe that the algorithms were not impacted by the imbalance of the data set (357 benign data points as compared to 212 malignant data points).

#### 2) Logistic Regression

Logistic regression, while a commonly used medical data classification algorithm, has performed the worst among all algorithms. This is due to the simplicity of the logistic regression model, which can only make a sigmoid curve decision boundary. While this sigmoid function increases the complexity of the decision boundary because it is nonlinear, it is not as complex as the decision boundaries created by random forest and deep learning classifiers.

#### 3) Random Forest

Random Forest’s performance should be interpreted cautiously, as it was determined that Random Forest was clearly and consistently overfitting. The algorithm was receiving perfect or near perfect results on the data the algorithm was trained on, while performing much worse (received scores about 5% worse) on testing data. This was a clear sign of overfitting. While many measures were taken to stop the algorithm’s tendency to overfit, it was eventually realized that the complexity of the model was too much for the WBCBD’s 569 data points. While the Random Forest model was created to both improve decision tree accuracies and stop the overfitting commonly associated with decision trees, the complexity of the decision boundaries that Random Forest created were too complex for this data.

#### 4) Linear Support Vector Classifier

Linear Support Vector Classifier (Linear SVC) performed significantly well, having the second highest testing score of 95.8%. The high classification score of Linear SVC is partially a result of the feature extraction model LDA. Because LDA’s goal is to make the data linearly separable, linear classification-based algorithms performed much better than they would without the use of LDA.

#### 5) Multilayer Perceptron

It was found that deep learning could not achieve high accuracies at predicting breast cancer malignancy from FNA biomarker. Instead, the algorithm performed relatively poorly, having the 4th best testing accuracy compared to the other algorithms tested. The relatively poor performance of deep learning can be attributed to the necessity to have a dropout layer after every regular layer of the network in order to avoid overfitting. However, dropout layers in neural networks significantly reduce the complexity of the decision boundary created by the neural network, causing it to fit the data with a model less superior than what deep learning usually does on datasets with millions of data points. Deep learning is known for requiring many data points to show a significant difference in testing accuracy as compared to conventional machine learning algorithms, and this project validates this knowledge, as deep learning did not show superior results to some of the more basic classifiers tested.

#### 6) Bagged Fuzzy-Rough Nearest Neighbors

Among all the ML algorithms evaluated, the novel ensemble algorithm, bagged fuzzy-rough nearest neighbors (BFRNN), was found to perform best in terms of all metric - testing accuracy, training accuracy and mAUC. The testing accuracy for this algorithm was the highest with the average value of 99.09%. Based on the performance of next best ML algorithm of Linear Support Vector Classifier, the novel algorithm BFRNN performed better by a margin of 3.5%. While Random Forest training accuracy (98.53%) was greater than BFRNN’s training accuracy (98.48%), the testing accuracy of Random Forest was much less, indicative of overfitting. The higher mAUC (1.0) observed with BFRNN suggests better classification characteristics of the algorithm. Onan [9] reported a similar high mAUC for his fuzzy-rough nearest neighbor classifier. With the novel BFRNN algorithm, feature selection and outlier removal steps were avoided making the implementation of the BFRNN algorithm much simpler.

## V. Conclusions

BFRNN has shown promise in classifying the malignant versus benign characteristics and this algorithm could be a useful tool in disease diagnosis. Many other useful dimensions such as patient medical history, BMI, images of cancer cells, common symptoms, etc can be considered while training in order to provide a holistic approach towards prediction of presence of active cancer.

## Data Availability

All data produced in the present study are available upon reasonable request to the authors

## Footnotes

1 With the BFRNN algorithm feature engineering and outlier detection methods specified in Figure 2 were not used. BFRNN uses Decision Tree Information Gain as the method for feature engineering. Outlier detection and removal were not performed.

## References

[1] L. Mellemkjær, S. Friis, J. H. Olsen, G. Scélo, K. Hemminki, E. Tracey, A. Andersen, D. H. Brewster, E. Pukkala, M. L. McBride et al., “Risk of second cancer among women with breast cancer,” International journal of cancer, vol. 118, no. 9, pp. 2285–2292, 2006.

[2] L. R. Borges, “Analysis of the wisconsin breast cancer dataset and machine learning for breast cancer detection,” Group, vol. 1, no. 369, pp. 15–19, 1989.

[3] S. Sakr, R. Elshawi, A. M. Ahmed, W. T. Qureshi, C. A. Brawner, S. J. Keteyian, M. J. Blaha, and M. H. Al-Mallah, “Comparison of machine learning techniques to predict all-cause mortality using fitness data: the henry ford exercise testing (fit) project,” BMC medical informatics and decision making, vol. 17, no. 1, pp. 1–15, 2017.

[4] W. Wolberg, O. Mangasarian, N. Street, and W. Street, “Breast Cancer Wisconsin (Diagnostic),” UCI Machine Learning Repository, 1995.

[5] D. S. Starnes, D. Yates, and D. S. Moore, The practice of statistics. Macmillan, 2010.

[6] L. Zadeh, “Fuzzy sets,” Information and Control, vol. 8, no. 3, pp. 338–353, 1965.

[7] Z.-H. Zhou and Y. Yu, “Adapt bagging to nearest neighbor classifiers,” Journal of Computer Science and Technology, vol. 20, no. 1, pp. 48–54, 2005.

[8] P. Emerson, “The original borda count and partial voting,” Social Choice and Welfare, vol. 40, pp. 353–358, 2013.

[9] A. Onan, “A fuzzy-rough nearest neighbor classifier combined with consistency-based subset evaluation and instance selection for automated diagnosis of breast cancer,” Expert Systems with Applications, vol. 42, no. 20, pp. 6844–6852, 2015.

